# Strategic Automated External Defibrillator Deployment Based on Geospatial Analysis of Historical Cardiac Arrest Locations

**DOI:** 10.1101/2025.10.03.25337296

**Authors:** Elizabeth Heckard, Harpriya Chugh, Katy Hadduck, Bryan F McNally, Ali Sovari, Jonathan Jui, Angelo Salvucci, Kyndaron Reinier, Sumeet S Chugh

## Abstract

**Objective:** We hypothesized that historical out-of-hospital sudden cardiac arrest (SCA) event locations can inform deployment of automated external defibrillators (AEDs).

**Methods:** Consecutive SCA events attended by EMS were ascertained prospectively in two identically designed community-based studies in California (Ventura County, SCA n=2,935) and Oregon (Multnomah County, SCA n=2,213). Clusters of ≥3 historical SCA events within a 100-meter radius were plotted using geospatial analysis. Proposed AED locations were positioned within 200-meters of the clusters. An existing AED registry in Ventura County (n=998) with coordinates was also plotted. Distances between SCAs and proposed AEDs were calculated.

**Results:** In Ventura County, 97 clusters of ≥3 SCAs were identified, covering 22% (n=653) of total SCAs with 90 proposed AEDs assigned to the clusters. The mean distance between individual SCA events and the nearest AED was 99.7 (SD=53.1) meters, representing a bystander AED response time of 3.0 (SD=1.6) minutes. Proposed bystander AED response times were significantly shorter than actual EMS response times [mean difference: 3.1 (95% CI: 2.9-3.4), p<0.0001]. Similar results were observed in Multnomah County. Compared to the existing AED registry in Ventura County, 18.9% (n=17) of our 90 proposed AEDs were within 200 meters of an existing AED. Of total SCAs in Ventura County,11.4% (n=334) were within 200 meters of an existing AED.

**Conclusions:** 16-22% of historical SCA events could potentially be covered by 78-90 AEDs. The remainder could be served by utilizing the standard EMS response. Outcomes studies that compare this geospatial event-localization strategy to current AED location strategies are warranted.

**Data Sharing statement:** The entire deidentified dataset, data dictionary and analytic code for this investigation are available upon request, from the date of article publication by contacting Sumeet S. Chugh, MD, at.

## Introduction

Out-of-hospital SCA affects approximately 360,000 individuals in the United States annually, with survival around 10%^[1]^. Bystander cardiopulmonary resuscitation (CPR) improves survival outcomes; however, in the United States, only 40% of bystanders perform CPR^[2]^. Deployment of AEDs also improves survival outcomes, but less than 10% of bystanders use AEDs^[3]^. For every minute an individual experiences ventricular arrhythmia (VA), a common occurrence in people suffering a SCA, their survival rate decreases by 7-10% without CPR and 3-4% with CPR; however, if an individual is defibrillated with an AED within 3 minutes of the onset of SCA, their survival rate is 25% higher than if they are defibrillated after 3 minutes (74% vs. 49%)^[3–5]^. One study based in Australia analyzed SCAs from 2010-2019 and reported that SCA events with bystander defibrillation using the closest AED displayed the highest survival rate (53%), even when compared to survival with EMS defibrillation by dispatched first responders (37%) and paramedics (28%)^[6]^.

There remains significant potential to optimize bystander and community use of AEDs for SCA. Some published studies have utilized historical SCA locations to identify opportunities to improve bystander AED response and deployment^[7, 8]^; however, these studies have relied on complete registries of existing AED locations, which are only available in certain US states. Efforts to improve AED registration in the U.S. are underway. PulsePoint is one organization looking to improve survival from SCA by providing public access to AED registries and encouraging participation by covering the registration fees of AEDs^[9]^. A state-wide AED registry was utilized in an Arizona study of SCAs occurring in public locations; however, they found a weak correlation between historical SCA events and physical AED locations, pointing to potential opportunities for optimizing AED placement^[7]^. Other studies have used historical SCA locations to evaluate methods to optimize EMS response through ambulances, mobile teams, volunteers, and drones^[10–16]^. A more calculated approach that focuses on the bystander and the AED as the heart of the community may be more impactful. We hypothesized that identifying clusters based on geospatial localization of historical SCA events could inform strategies to improve placement and deployment of AEDs. We used both Euclidean and Walking-line distances to calculate a more realistic range of potential bystander response times, facilitating a better evaluation of the potential impact of AEDs on survival for each individual SCA.

## Methods

### Study design and setting

Consecutive events of out-of-hospital SCA attended by EMS were ascertained prospectively in two ongoing population-based studies in the United States, PRESTO (Prediction of Sudden Death in Multi-Ethnic Communities) and SUDS (Sudden Unexpected Death Study)^[17, 18]^. PRESTO, based in Ventura County, California (population ∼850k), and SUDS, based in Multnomah County, Oregon (population ∼800k), follow identical study designs. Ventura County has a population density of 458.4 individuals per square mile, and Multnomah County has a population density of 1,891.9 individuals per square mile^[19, 20]^. We conducted evaluations of SCA event clusters and potential AED locations separately in the two populations to assess the consistency of our findings in separate geographical regions.

### Selection of participants

SCA events were identified by each county’s EMS system, and adjudication was performed by trained research personnel. SCA was defined as the sudden loss of pulse of likely cardiac origin^[1]^. Events of non-cardiac origin (e.g. substance abuse, trauma, or terminal illness) were excluded. The PRESTO study analysis included 2,935 SCA events (Age: 21 d-103 yrs) with resuscitation attempted by EMS personnel between February 1st, 2015-January 31st, 2023. The SUDS analysis included 2,213 SCA events (Age: 19 d-111 yrs) with resuscitation attempted by EMS personnel between February 1st, 2012-January 31st, 2020. Patient information obtained from regional EMS systems included SCA location (address, city, state, zip code), EMS response time, time of 9-1-1 call, and date of SCA. Both studies were approved by the Institutional Review Boards (IRB) of Ventura County Medical Center, Oregon Health and Science University, Cedars-Sinai Health System, and all other participating hospitals and health systems. Written informed consent was required for SCA survivors.

### Definition of SCA clusters

The research question focused on strategic proposed AED deployment based on identifying geospatial clusters of historical SCA locations. First, we evaluated different parameters and conducted sensitivity analyses to define our SCA clusters. Our main independent variables were 1) Distance between SCAs within a cluster (75, 100, and 125 meters) and 2) Number of SCAs per cluster (2, 3, 4, 5). Our goal was to minimize bystander response time and number of clusters (thus reducing the number of AEDs required) and maximize the proportion of SCAs covered by the proposed AED locations. We defined SCA Clusters as ≥3 SCA events within a 100-meter radius. Our clusters of ≥3 SCA events within 100 meters minimized the distance between SCA events, proposed bystander response time, and the number of clusters, and maximized the proportion of SCA events covered. Second, proposed AED locations (AEDs at fixed locations within walking distance), were plotted <200 meters from each SCA in a cluster. Per the American Heart Association (AHA) guidelines it takes 1.5 minutes to walk 100 meters^[2]^. Previous research shows that AEDs are most effective if applied within 3-5 minutes of the SCA, and we defined AED distances based on this recommended response time frame ^[3–5]^.

A voluntary registry of public-location AEDs maintained by Ventura County EMS Services (n=998 AEDs) was used to compare the location of our proposed AEDs to existing AEDs in Ventura County. This registry did not include many schools, care facilities, and other buildings which require an AED on-site under California law, and thus was incomplete; however, publicly available complete AED registries were not available for either Ventura or Multnomah counties. Proposed AEDs were deemed as “covered” by an existing AED if they were in the same or neighboring building or <300 meters.

### Geospatial analysis

SCA locations (address, city, state, zip code) were entered in QGIS Desktop 3.23.3 for geospatial analysis using the MMQGIS plug-in^[21–23]^. Geocoded points (latitude, longitude) were output onto the map in Geopackage or shp files. Incorrectly geocoded points were corrected by crosschecking against GoogleMaps and re-entered into QGIS for analysis. From here, the Density-Based Spatial Clustering of Application with Noise (DBSCAN) tool generated clusters of SCA location points grouped by ≥3 SCA events within a 100-meter radius^[22, 23]^. DBSCAN uses Euclidean distances to generate clusters within a certain radius, calculated as straight-line distances between two points. This methodology can be less practical when estimating bystander walking time. Therefore, SCA events were also mapped to the nearest AED using the QuickOSM query feature inputting highways, roads, and sidewalks as walking routes. This allowed for a more realistic range of potential bystander response time between a SCA event and the nearest AED. The QNEAT3 plugin (OD Matrix from Layers as Lines m:n) then generated walking-line distances in meters. K-means, hierarchal, and UMAP clustering methods were also evaluated against DBSCAN; however, both K-means and hierarchal clustering require specifying the number of clusters beforehand, while our goal was to determine the number of clusters using historical data, and UMAP is better suited for visualizing data in 2D or 3D formats.

The OpenStreetMap tool positioned proposed point-of-care AED locations on existing buildings or structures. Based on findings from other studies and randomized controlled trials (RCTs), a feasible distance between a SCA location and an AED is <200 meters^[24, 25]^. While the AHA recommends positioning an AED within 100-meters of a SCA event,^[2]^ we set the maximum distance between each SCA event in a cluster and the nearest proposed AED location to 200 meters, assuming the mean distance would be less (Figure 1). For each SCA event, the potential bystander AED response time was estimated as roundtrip walking time from the exact geocoded location of that SCA event to the proposed AED location and back (∼1.5 minutes per 100m). An AED could potentially cover multiple SCA clusters if it is positioned within 200-meters of every SCA event in those clusters. Non-residential public buildings (Ex. school, grocery store, gym, church) were prioritized for AED placement, with the goal of broader accessibility.

**Figure 1).**
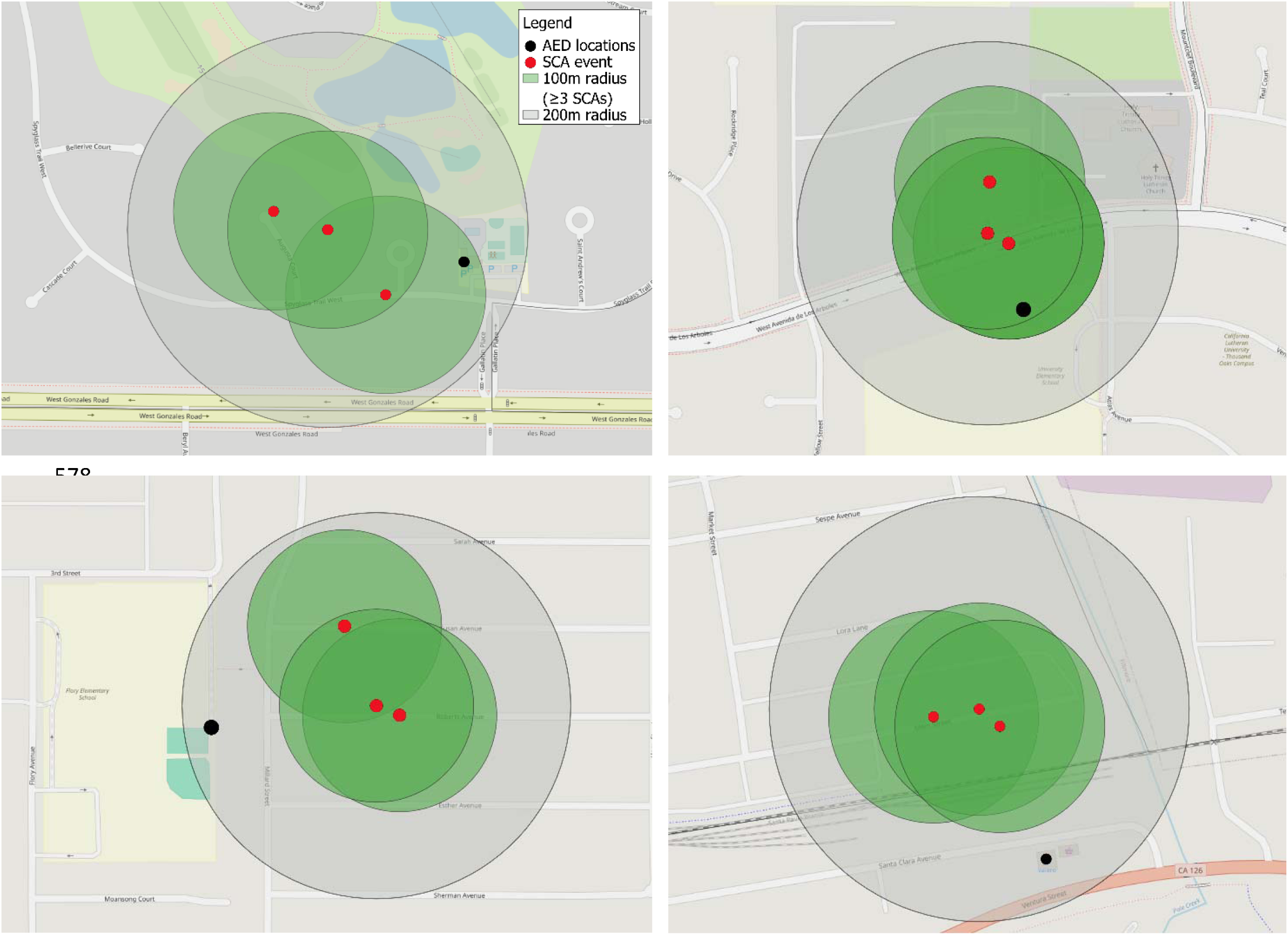
Examples of clusters of ≥3 SCA events within a 100-meter radius created on QGIS Desktop 3.23.3. Proposed AED locations were placed within 200 meters of each SCA event.

Visualization of the spatial clustering model was examined using the Heatmap Kernel Density estimation algorithm in QGIS. We grouped 8-year SCA data for Ventura County (2015-2023) first, and Multnomah County (2012-2020) second (Figure 2). The maps illustrate the density of each cluster by the number of SCA events.

**Figure 2).**
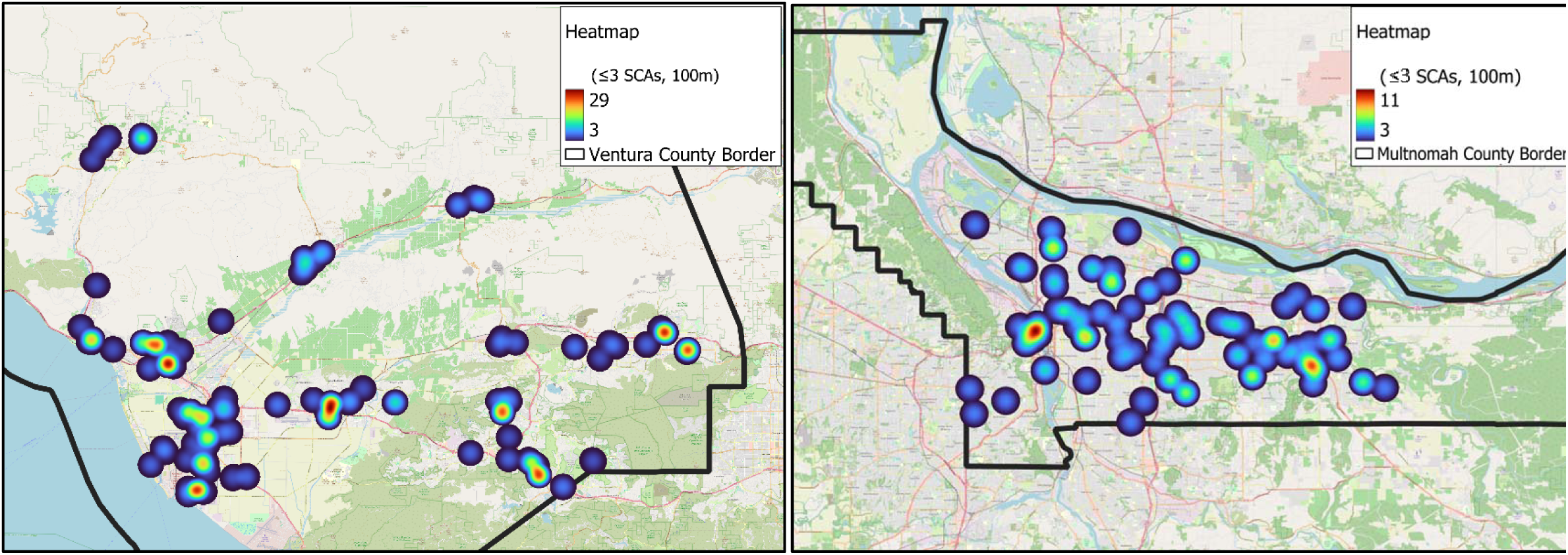
SCA density clusters in Ventura and Multnomah County from QGIS Desktop 3.23.3.

### Evaluating SCA locations over time

The Spatial-Temporal DBSCAN (ST-DBSCAN) tool expands from the DBSCAN tool to include a moving time period in addition to generating clusters^[22–23]^. We analyzed potential temporal changes in cluster patterns to evaluate whether historical SCA locations are stable over time and thus could inform future placement of AEDs. A moving time period implies that every 4-year period would be evaluated (PRESTO: 2015-2019, 2016-2020, 2017-2021, 2018-2022, 2019-2023; SUDS: 2012-2016, 2013-2017, 2014-2018, 2015-2019, 2016-2020). There is one output, like DBSCAN, which acts as an average across the moving time periods (Table S2).

### Statistical analysis

Demographics and descriptive characteristics of individuals with SCA in the Ventura and Multnomah studies were presented as frequencies and percentages for categorical variables and mean ± SD for continuous variables. Overall average response times were presented as mean ± SD values. Paired t-tests were used to evaluate individual differences between EMS response time and proposed (Euclidean) bystander AED response time and presented as mean difference (95% CI) and p-values.

## Results

### Characteristics of study subjects

From the PRESTO study, we identified 3,029 SCA events in Ventura County from 2015-2023 after detailed adjudication. Of these events, 94 (3%) were excluded because the SCA locations could not be geocoded due to incorrect addresses (n=2 excluded due to their location being outside of Ventura County map boundaries). A total of 2,935 SCAs were included in the final analysis. From the SUDS study, we identified 2,543 SCA events in Multnomah County from 2012-2020 after detailed adjudication and exclusion. Of these events, 330 (13%) SCA locations could not be geocoded due to incorrect addresses (n=4 excluded due to their location being outside of Multnomah County map boundaries). 2,213 SCAs were included in the final analysis.

Table 1 shows the demographics and circumstances of SCA in PRESTO and SUDS. Most events from both counties were male (Ventura: n=1,923, 66%; Multnomah: n=1,485, 67%), White, non-Hispanic (Ventura: n=1,746, 60%; Multnomah: n=1,485, 67%) and older (Ventura: 70 yrs. +/- 16; Multnomah: 64 yrs. +/- 18). The most common SCA location for both studies was home (Ventura: n=2,199, 75%; Multnomah: n=1,410, 64%), followed by care facility (Ventura: n=372, 13%; Multnomah: n=273, 12%). About half of SCAs had bystander CPR administered (Ventura: n=1,585, 54%; Multnomah: n=1,161, 53%) and were witnessed (Ventura: n=1,418, 48.3%; Multnomah: n=1,204, 54.4%). Few had bystander AED use (Ventura: n=92, 3%; Multnomah: n=121, 6%). The most common presenting rhythm was Asystole (Ventura: n=1,414, 48.2%; Multnomah: n=913, 41.3%), followed by VFVT (Ventura: n=704, 24.0%; Multnomah: n=750, 33.9%) and PEA (Ventura: n=795, 27.1%; Multnomah: n=485, 21.9%). The average EMS response time reported across total SCA events was 6.3 minutes (SD=3.0) in Ventura County and 7.4 minutes (SD=3.7) for Multnomah County.

**Table 1).** Demographics and arrest circumstances among SCA events in the Ventura and Multnomah County study populations.

|  | Ventura County<br>2015-2023<br>(n=2,935) | Multnomah County<br>2012-2020<br>(n=2,213) |
| --- | --- | --- |
| Gender<sup>a</sup> |  |  |
| Male | 1,923 (65.5%) | 1,485 (67.1%) |
| Age (mean, SD) | 70.4 ± 16.4 | 64.2 ± 17.8 |
| Race/ethnicity<sup>b</sup> |  |  |
| White | 1,746 (59.5%) | 1,640 (74.1%) |
| Hispanic | 766 (26.1%) | 77 (3.5%) |
| Asian | 188 (6.4%) | 103 (4.7%) |
| Black/African American | 67 (2.3%) | 252 (11.4%) |
| American Indian/Alaska Native | 12 (0.4%) | 21 (0.9%) |
| Native Hawaiian/Pacific Islander | 12 (0.4%) | 21 (0.9%) |
| Other | 9 (0.3%) | 10 (0.5%) |
| SCA Location |  |  |
| Home | 2,199 (74.9%) | 1,410 (63.7%) |
| Care Facility | 372 (12.7%) | 273 (12.3%) |
| Public, outside | 203 (6.9%) | 273 (12.3%) |
| Public Bldg. or Facility | 161 (5.5%) | 257 (11.7%) |
| Witnessed<sup>d</sup> |  |  |
| Yes | 1,419 (48.3%) | 1,204 (54.4%) |
| Bystander CPR |  |  |
| Yes | 1,585 (54.0%) | 1,161 (52.5%) |
| Initial Rhythm<sup>e</sup> |  |  |
| VFVT | 704 (24.0%) | 750 (33.9%) |
| Asystole | 1,414 (48.2%) | 913 (41.3%) |
| PEA | 795 (27.1%) | 485 (21.9%) |
| Other | 19 (0.6%) | 39 (1.8%) |
| Bystander AED |  |  |
| Yes | 92 (3.1%) | 121 (5.5%) |
| EMS Response Time (mean, SD)<sup>f</sup> | 6.3 ± 3.0 | 7.4 ± 3.7 |

<sup>a</sup>n=1, missing
<sup>b</sup>n=224, missing
<sup>d</sup>n=7, missing
<sup>e</sup>n=29, missing
<sup>f</sup>n=3, missing

### Proposed AED Response Strategy

Of the total SCAs in Ventura County, 22% (n=653) were grouped into clusters of ≥3 SCAs within a 100-meter radius. We report that by increasing the cluster radius distance from 75 to 100 meters, we can cover an additional 40-67 SCA events with an additional 10-18 AEDs, increasing coverage rate by 10-11%. This is based upon using a cluster size of 3 SCAs per cluster. 97 clusters of ≥3 SCAs (mean 14.0 SCA events per cluster, range 3-29) were created (Figure 3). 90 proposed AED locations were required to cover the 97 clusters. Of these 90 proposed AED locations, most were assigned to non-residential, public buildings (Non-residential n=72, 80%; Residential n=18, 20%). The overall mean Euclidean distance between an SCA event in a cluster and a proposed AED was 99.7 meters (SD=53.0), and the mean Walking-line distance was 136.7 meters (SD=115.9) (Table 2). In Multnomah County, 16% (n=353) of total SCAs were grouped into 83 clusters of ≥3 SCAs (mean 5.1 SCA events per cluster, range 3-11). 78 proposed AED locations were required to cover the 83 clusters; most were assigned to non-residential, public buildings (Non-residential n=63, 81%; Residential n=15, 19%). The overall mean Euclidean distance between an SCA event in a cluster and a proposed AED was 84.1 meters (SD=47.8), and the mean Walking-line distance was 130.0 meters (SD=134.2).

**Figure 3).**
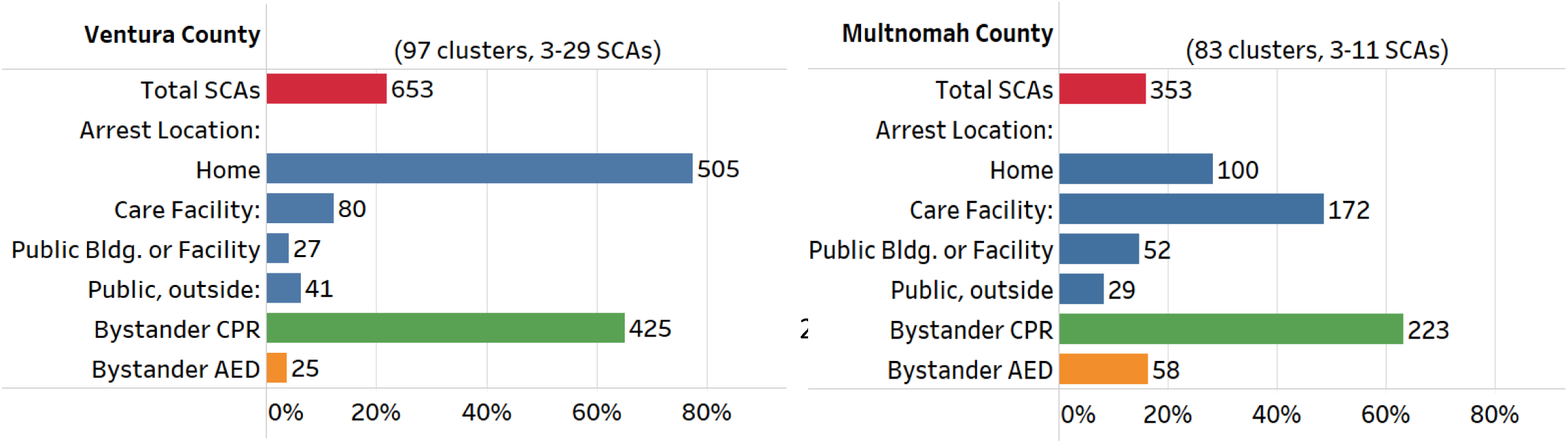
SCA descriptive characteristics for Ventura and Multnomah County.

**Table 2).**
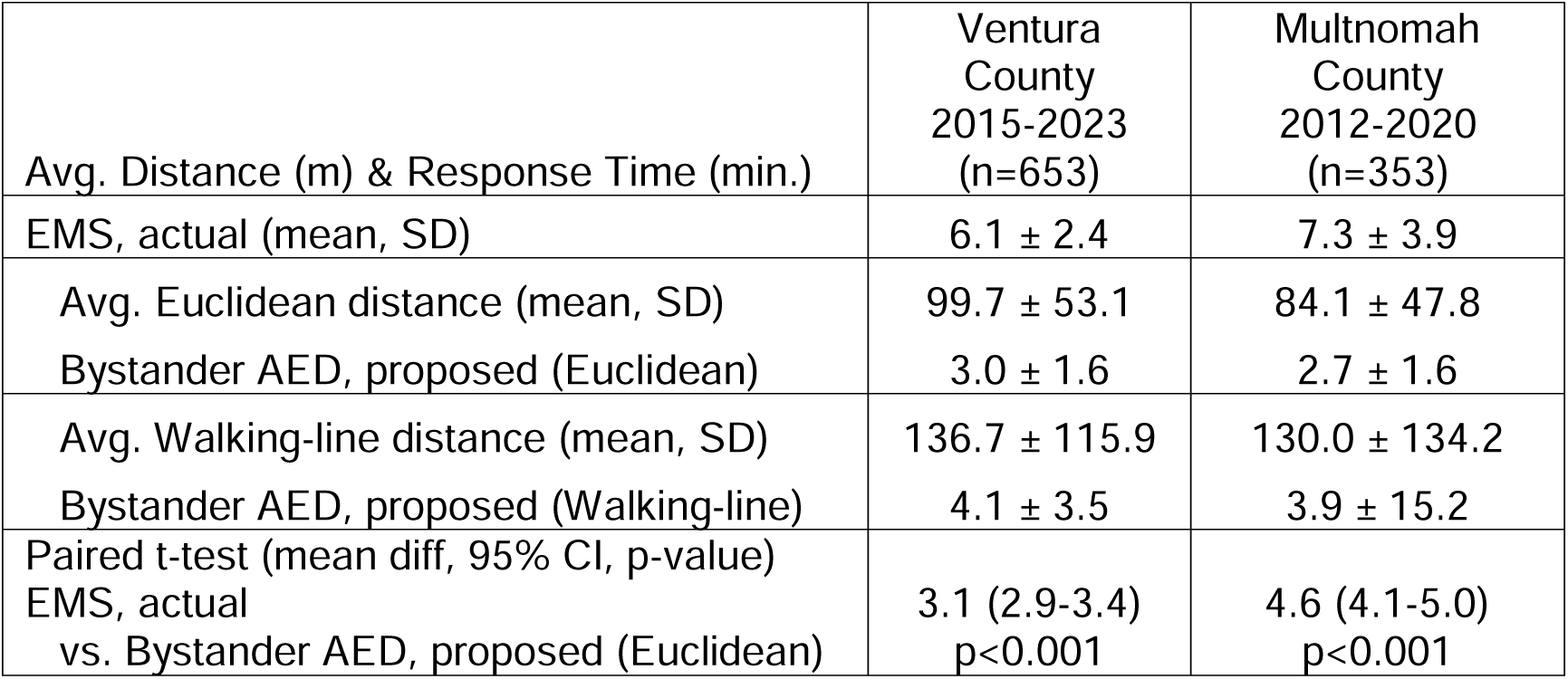
Average Euclidean and walking-line distances between SCA clusters and the nearest proposed AED for Ventura and Multnomah County. Overall average actual and proposed Euclidean and Walking-line response times. Paired t-test values (mean difference, 95% CI, p-value) between actual and proposed (Euclidean) bystander response time for Ventura and Multnomah County.

Table 1) Demographics and arrest circumstances among SCA events in the Ventura and Multnomah County study populations.
|  | Ventura<br>County<br>2015-2023<br>(n=653) | Multnomah<br>County<br>2012-2020<br>(n=353) |
| --- | --- | --- |
| Avg. Distance (m) & Response Time (min.) |  |  |
| EMS, actual (mean, SD) | 6.1 ± 2.4 | 7.3 ± 3.9 |
| Avg. Euclidean distance (mean, SD) | 99.7 ± 53.1 | 84.1 ± 47.8 |
| Bystander AED, proposed (Euclidean) | 3.0 ± 1.6 | 2.7 ± 1.6 |
| Avg. Walking-line distance (mean, SD) | 136.7 ± 115.9 | 130.0 ± 134.2 |
| Bystander AED, proposed (Walking-line) | 4.1 ± 3.5 | 3.9 ± 15.2 |
| Paired t-test (mean diff, 95% CI, p-value) |  |  |
| EMS, actual<br>vs. Bystander AED, proposed (Euclidean) | 3.1 (2.9-3.4)<br>p<0.001 | 4.6 (4.1-5.0)<br>p<0.001 |

Of our proposed AEDs, 18.9% (n=17) were within 200 meters of an existing AED in the registry, meaning that 81.1% (n=73) of our proposed AEDs were outside 200 meters. The overlapping AEDs were in schools mainly, and a community center and a medical building. Only 11.4% (n=334) of total SCAs were within 200 meters of an existing AED, meaning that 88.6% (n=2,601) were outside of 200 meters to the closest existing AED. In addition, a greater proportion of our proposed AEDs were residential (28%, n=25) compared to existing AEDs (6%, n=60).

### Potential Improvements in Response Time & Survival

Overall average times for EMS and Bystander response are shown in Table 2. For Ventura County, the average Euclidean distance was 99.7 meters (SD=53.1), with a corresponding average bystander response time of 3.0 minutes (SD=1.6). Average Walking-line distance was 136.7 meters (SD=115.9), with a corresponding average bystander response time of 4.1 minutes (SD=3.5). For Multnomah County, the average Euclidean distance was 84.1 meters (SD=47.8), with a corresponding average bystander response time of 2.7 minutes (SD=1.6). Average Walking-line distance was 130.0 meters (SD=134.2), with a corresponding average bystander response time of 3.9 minutes (SD=15.2). Proposed bystander AED response time based on Euclidean distances showed a benefit of 3.1 minutes saved in Ventura County, and 4.6 minutes saved in Multnomah County compared to EMS response time. Walking-line distances showed a benefit of 2 minutes saved in Ventura County, and 3.4 minutes saved in Multnomah County compared to EMS response time. Mean difference, 95% confidence interval, and statistically significant p-values were calculated for both Ventura (mean diff. 3.1, 95% CI: 2.9-3.4, p<0.0001) and Multnomah County (mean diff. 4.6 95% CI: 5.9-7.1, p<0.0001).

### Spatial-Temporal Analysis

Across Ventura and Multnomah County, the number of SCA events, clusters, cluster size, and average distance to the nearest AED stayed relatively consistent with our original findings using 4-year moving time periods (Table S2). The spatial-temporal analysis produced clusters that covered between 84-94% of the SCA events in the original clusters [Ventura: 4-years (93%); Multnomah: 4-years (90%)]. More importantly, average distance to the nearest AED was consistent, with the average distance to the nearest AED ranging from 97.7-99.7 meters for Ventura County, and 81.3-84.1 meters for Multnomah County.

## Discussion

Among 2,935 SCA events in Ventura County from 2015-2023, we used geospatial mapping to identify 97 clusters of ≥3 SCA events within a 100-meter radius (n=653 events, 22% of total SCAs). We estimated that these 653 SCA events could have been covered by 90 proposed AEDs within 200 meters of each SCA. In a separate population, among 2,213 SCA events in Multnomah County from 2012-2020, we identified 83 clusters of ≥3 SCA events (n=353 events, 18% of total SCAs) that could have been covered by 78 proposed AEDs. The average distance range between each SCA event and the nearest proposed AED was 99.7-136.7 meters for Ventura County, and 84.1-130.0 meters for Multnomah County, corresponding to an average proposed bystander roundtrip response time between 3.0-4.1 minutes for Ventura County, and 2.7-3.9 minutes for Multnomah County for retrieval of an AED. Comparing proposed bystander AED response times to actual EMS response times, time to defibrillation could be reduced by 3.1-4.6 minutes across both counties through AED placement optimization. Of our proposed AEDs, 18.9% (n=17) were within 200 meters of an existing AED in the registry, meaning that 81.1% (n=73) of our proposed AEDs were outside 200 meters. Only 11.4% (n=334) of total SCAs were within 200 meters of an existing AED, meaning that 88.6% (n=2,601) were outside of 200 meters to the closest existing AED. Bystander AED use was low in both Ventura and Multnomah County (3.1-5.5%) and this could further demonstrate the need for optimization of AED locations. Analysis of temporal patterns showed relatively stable SCA event locations over time. This may demonstrate the reliability of historical SCA locations in planning future AED deployment.

Among studies performed in the United States, this analytical, community-based geospatial approach is novel. Outside of the US, some published studies have proposed AED placement utilizing historical SCA locations and AED registry data. Researchers in Switzerland defined their SCA hotspots as ≥5 historical SCAs within 100-meters. By placing theoretical AEDs in their 40 hotspots, 225 historical SCAs would be additionally covered, increasing the coverage rate by 10%^[11]^. A second study from Denmark used similar definitions for AED coverage of SCA events^[26]^. This study did not create proposed AED locations but rather relied on the 552 registered AEDs in the Danish AED Network. 537 SCA events (29%) were covered by an AED. Karlsson et al. also utilized the Danish AED registry but expanded AED coverage to <200-meters, covering 566 SCA events^[25]^. In both study regions, we conducted similar analyses. In an Australian study, AEDs within 100-meters of an SCA event were only available for 4% of SCAs that occurred at home and 23% of SCAs that occurred in public areas; when extended to 200-meters, AED availability increased by 8-18% between homes and public areas^[6]^. For Ventura County, a greater proportion of proposed AEDs that were not covered by existing AEDs were residential (28%) compared to covered AEDs (6%), showing the need for residential SCA events in AED optimization analyses. Karlsson et al. revealed that SCAs within 200-meters of an AED were 9% more likely to receive bystander CPR or AED response, and 12% more likely to achieve 30-day survival than SCAs outside 200-meters^[25]^. In both study regions, we report that bystander CPR was performed during 53-54% of SCA events and bystander AEDs were used for 3-6% of SCA events.

The lack of comprehensive information regarding AED locations and accessibility creates a substantial barrier to AED use in the US. Firstly, although PulsePoint AED, a public non-profit organization, is building an emergency AED registry that is available free of charge in the US, there are currently no comprehensive, national AED registries^[9]^. For our purposes we used an incomplete AED registry in Ventura County to compare to our proposed AED locations because a more complete registry was not publicly available. This inconsistency in registries makes it difficult for an EMS dispatcher or even a bystander to locate an AED. In addition, shockable rhythms are highest (79%) in individuals experiencing a SCA in public locations when bystander AED is administered, and this percentage drops drastically in residential locations with or without bystander AED use (36%, 25%)^[27–28]^.Secondly, updating AED registries (location, device/battery) is voluntary, so complete information regarding locations and accessibility of public AEDs is unknown. Thirdly, there are limited requirements for AEDs in public areas outside of airports, schools, and fitness centers, though these also vary by state. In Oregon, AEDs are required in schools, colleges, health clubs, large occupancy environments, camps, and swimming pools, whereas in California, AEDs are required in health clubs, large occupancy buildings, public swimming pools, schools with interscholastic programs, and specific commuter trains^[29, 30]^. Many of the proposed AEDs in our study were positioned in similar buildings like schools and gyms, but we also prioritized public buildings with extensive hours for accessibility like religious buildings or grocery and convenience stores.

For SCAs that fell outside our previous parameters of ≥3 SCA events within a 100-meter radius, we hypothesize that the community and CPR-trained volunteers could complement the standard EMS response. Traditionally, studies in Sweden have recruited CPR-trained civilian first responders by phone to aid before EMS arrival^[31]^. In addition, SCA events falling in rural areas or larger distances could benefit from novel strategies in drone-assisted deliveries. In these rural areas, EMS response times are exceedingly high (avg. 20-26 min.)^[31, 32]^. Many pilot studies have already implemented this practice in their models where distances extend beyond >1 mile ^[13–15]^.In the US, Starks et al. investigated drone-assisted AEDs to decrease response time for SCA victims when distances are too far for the bystander to walk or traditional EMS to drive^[34]^. Studies on the drone delivery of AEDs compared to EMS services have shown benefits in response time ranging from 3.2-16.7 minutes over mean distances of 2 miles^[13–15]^. In Europe, Claesson et al. performed a pilot study of 18 drones. With a median flight distance of 3.2 km (2 miles), the time from dispatch to arrival was 5.2 minutes for drone-assisted AED delivery and 22 minutes for EMS response^[13]^. Schierbeck and colleagues followed up this study by employing drones in the first real-world evaluation of drone-assisted AED delivery in Sweden. They showed a median time-benefit of 3.2 minutes in drone-delivery compared to ambulance arrival; however, weather, air traffic operating hours, and accuracy of intended site drop-off were presented as potential obstacles for replication^[14, 15]^.

## Strengths and Limitations

The Ventura and Multnomah County populations are similar in size and SCA incidence, and replication in a second study population to evaluate consistency in clustering is a strength of our study. However, since our clustering methodology relied on historical SCA events, the deployment model may vary in other communities based on location, size of the US County, and other characteristics. For example, in Multnomah County, a larger proportion of SCA events occurred in care facilities, likely explained by facility concentration pockets in specific parts of the County. Though we were able to confirm our geocoded results from QGIS in Google Maps, we were not able to obtain latitude and longitude values for 3% of PRESTO events and 13% of SUDS events due to unconfirmed addresses. Unfortunately, in a perfect scenario we would have access to a complete, updated AED registry for both Ventura and Multnomah County, but this was not publicly accessible. AEDs are not accessible 24/7 and they are not helpful for non-shockable rhythm; however, we tried to optimize proposed AED locations within public buildings to increase hours of accessibility. Lastly, EMS response time does not reflect the total time between SCA and the beginning of resuscitation efforts, due to the difference between collapse time and the time the ambulance leaves the station, and time between EMS arrival and defibrillation. We may be overestimating the utility of the ambulance and underestimating the total response time. If so, the proposed bystander AED response time could have a larger time benefit than anticipated.

## Conclusion

In Multnomah and Ventura Counties, 16-22% of historical SCA events occurring between 2012-2023 could be covered by 78-90 proposed AED locations, with potential improvements in the time between collapse and defibrillation of 3.1-4.6 minutes compared to EMS response times. These findings provide a theoretical construct and rationale for future studies aiming to optimize bystander AED utilization at the US county level.

## Data Availability

All data produced in the present study are available upon reasonable request to the authors.

---

**Supplementary Table 1).**
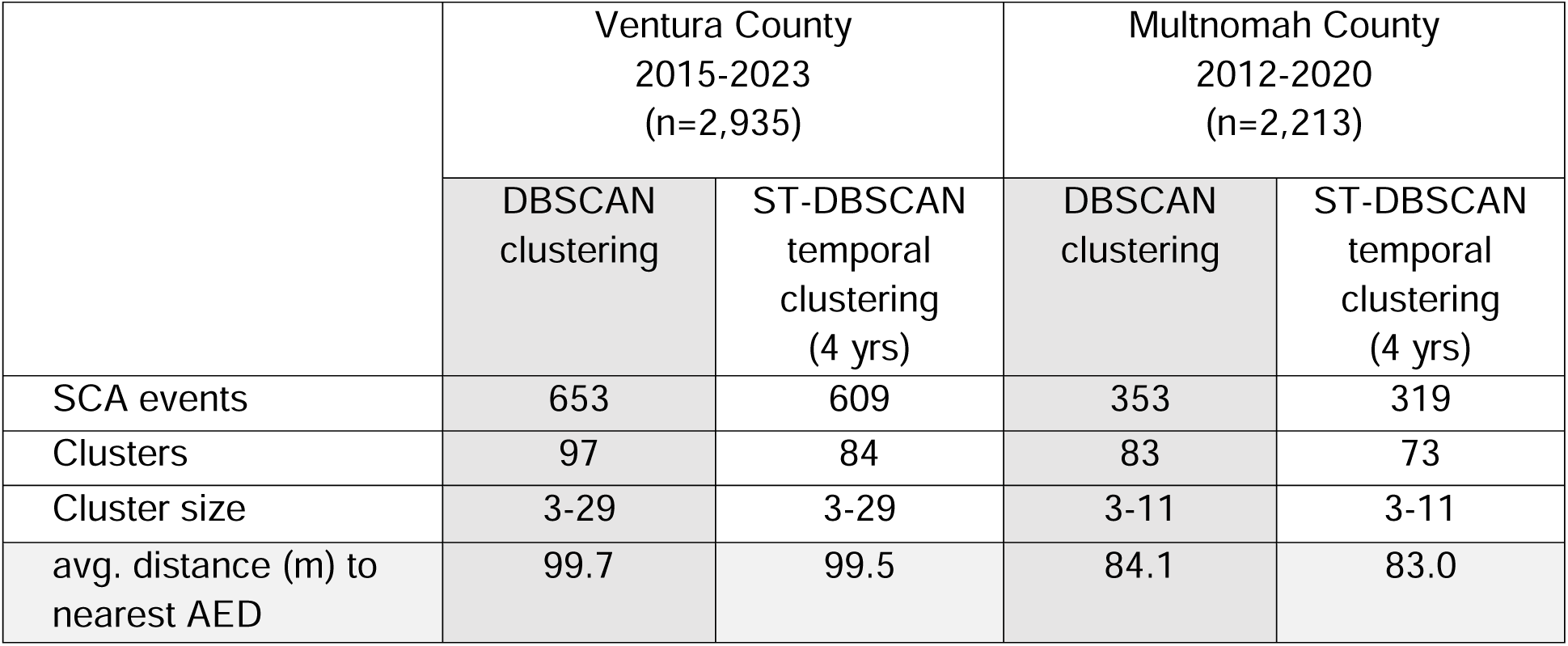
Adjusting for temporal changes in SCA event clustering (Euiclidean distances) for Ventura and Multnomah County.^†^ †The Spatial-Temporal DBSCAN (ST-DBSCAN) tool from QGIS generates clusters based on minimum number of points per cluster, maximum allowed distance between points, and a moving time period^[27]^. This methodology accounts for temporal changes in cluster patterns and can evaluate whether clusters are stable over time. A moving time period of 4 years for example, implies that every 4-year period would be evaluated in the datasets. (PRESTO: 2015-2023 total; 2015-2019, 2016-2020, 2017-2021, 2018-2022, 2019-2023; SUDS: 2012-2020 total; 2012-2016, 2013-2017, 2014-2018, 2015-2019, 2016-2020). This produces one output which is an average across all moving time periods, shown above.

## Notes

### Competing Interest Statement

The authors have declared no competing interest.

### Funding Statement

This study did not receive any funding.

### Author Declarations

Both studies were approved by the Institutional Review Boards (IRB) of Ventura County Medical Center, Oregon Health and Science University, Cedars-Sinai Health System, and all other participating hospitals and health systems. Written informed consent was required for SCA survivors.

